# “There’s a big tag on my head”: exploring barriers to treatment seeking with women who use methamphetamine in Sydney, Australia

**DOI:** 10.1101/2022.08.01.22278295

**Authors:** Brendan Clifford, Kate Van Gordon, Fiona Magee, Victoria Malone, Krista Siefried, Duncan Graham, Nadine Ezard

## Abstract

**Background:** Australia has a high prevalence of regular use of methamphetamine (MA). While half of people who use MA regularly are women, they make up only one third of people seeking treatment for MA use disorder (MaUD). There is a lack of qualitative research into the facilitators and barriers to treatment for women who use MA regularly. The study seeks a better understanding of the experiences and treatment preferences of women who use MA, to inform person-centred changes in practice and policy that break down barriers to treatment.

**Methods:** We conducted semi-structured interviews with 11 women who frequently use MA (at least once a week), and who are not engaged in treatment. Women were recruited from health services surrounding a stimulant treatment centre at an inner-city hospital. Participants were asked about their MA use and health service needs and preferences. Thematic analysis was completed using Nvivo^®^ software.

**Results:** Three themes were developed from participants’ responses around experiences of regular MA use and treatment needs: 1. Resistance of stigmatised identity including dependence; 2. Interpersonal violence; 3. Institutionalised stigma. Clear service delivery preferences were also elicited, including continuity of care, integrated health care, and provision of non-judgmental services.

**Conclusion:** Gender-inclusive health care services for people who use MA should actively work to address stigma, support a relational approach to assessment and treatment, and seek to provide structurally competent health care that is trauma and violence informed, and integrated with other services. Findings may also have application for substance use disorders other MA.

## Background

The use of MA is a growing public health concern globally (Farrell et al., 2019), with MA related death rates increasing in Australia from 0.14 to 0.44 per 100,000 population between 2012 and 2016 (Man et al., 2022). Harms associated with MA use include depression, anxiety, psychosis, cardiovascular disease, cerebrovascular disease (Chomchai & Chomchai, 2015), and the development of MaUD (Degenhardt et al., 2017). Evidence-based therapies for MaUD include psychosocial interventions such as Cognitive Behavioural Therapy (CBT) (Asharani et al., 2020) with ongoing work to develop adjunct pharmacotherapies (Siefried, Acheson, Lintzeris, & Ezard, 2020). In Australia, despite high rates of regular use of MA and MaUD, there is relatively low treatment utilisation, with a gap of five (Lee, Harney, & Pennay, 2012) to ten years (Brecht, Lovinger, Herbeck, & Urada, 2013) between first problematic use of MA and the seeking of treatment. This is partly due to self-perceived non-problematic use, even after people have begun to experience harms associated with MA (Quinn, Stoove, Papanastasiou, & Dietze, 2013). People who use MA, however, also report low confidence in the efficacy of treatment services and cite the opiate-centric nature of alcohol and other drug (AOD) services as barriers to help-seeking (Cumming, Troeung, Young, Kelty, & Preen, 2016; Kenny, Harney, Lee, & Pennay, 2011; Rapp et al., 2006). Specialist treatment services that provide psychological and medical interventions for people experiencing MaUD delivered within a non-judgmental, harm reduction framework have been found to be attractive to participants (Brener et al., 2018) and effective in providing significant reductions in MA use and associated harms (McKetin et al., 2013).

Substance use remains deeply gendered in Australia, being seen as of rite of passage and a form of active exploration in masculinity, but linked with “ineffective coping mechanisms” (Frydenberg, 2014) and a need to quell internal psychological struggles when associated with femininity (Keane, 2017). Importantly, evidence suggests that once in treatment for MaUD, outcomes for women may be better than those for men. In a longitudinal Californian study of people in treatment, for instance, women demonstrated greater improvement in family relationships and medical problems compared to men. This was despite women in the sample being more likely than men to be unemployed, have childcare responsibilities, live with someone who also used substances, have been physically or sexually abused, and to have more psychiatric symptoms (Hser, Evans, & Huang, 2005). In Australia, however, women are less likely to participate in treatment (McKetin & Kelly, 2007), making up only a third of people who access treatment for MaUD (Australian Institute of Health and Welfare, 2020), despite being as likely as men to use MA weekly (Roche, McEntee, Fischer, & Kostadinov, 2015).

This study was undertaken to understand the nature of barriers to specialist MaUD treatment for women in metropolitan area of Sydney, Australia. Rather a fixed binary epidemiological or clinical category, it understands gender as complex social phenomenon that extends beyond these classifications (Martin & Aston, 2014), and seeks to build the voices of women into contemporary clinical practice.

## Methods

### Design

An exploratory qualitative methodology was used to assess experiences of women who regularly MA but who have not accessed a specialised MA treatment service. The study setting was an inner-city tertiary Australian hospital with a free specialist treatment program for people with that provides assessment and early intervention (Brener et al., 2018) and outpatient counselling (McKetin et al., 2013). Ethics approval was obtained from St Vincent’s Hospital Human Research Ethics Committee (LNR/15/SVH/469).

### Sampling and data collection

Study advertisements were placed in local and community health centres, drug and alcohol services, and women’s refuges seeking interviews with people who identified as women and who used MA at least weekly, were not engaged in MA specific treatment, and were aged at least 18 years. Prospective participants were asked to contact the investigator team by telephone to arrange an interview time, where informed consent was obtained. Participants were given an AU$20 grocery voucher prior to completing demographic and substance use questionnaires, the Severity of Dependence Scale (SDS) (Gossop et al., 1995) and undertaking a semi-structured interview exploring participants experiences of methamphetamine use, health care services and treatment preferences for MaUD.

### Analysis

The interview data are recognised as being co-created by participant and interviewer, and both the data and its analysis influenced by the perspectives and genders of the researchers (Broom, Hand, & Tovey, 2009). The interviews were transcribed, and analysed thematically (Braun & Clarke, 2006). Authors KVG, FM, BC and VM initially coded two transcripts each using NVivo^®^ (QSR International) software examining participants’ relationship to and experiences of MA use; barriers experienced in access to care; use of health services; and perceived treatment needs. The study team then met to examine emergent subthemes from the codes, exploring linkages between themes and subthemes, searching for negative or deviant examples, extracting quotes to exemplify arguments, and developed a thematic framework with which to analyse the remaining transcripts. After the remaining transcripts were coded, the team met again to re-evaluate the validity of the themes and draw out meta-themes.

## Results

A total of eleven women who used methamphetamine were interviewed, mainly recruited from non-gender specific AOD services. Participant characteristics are presented in **Table 1**. Four themes were developed from the data: identity, stigma and resistance; interpersonal violence; institutionalised stigma; and service delivery preferences. Quotes are provided to illustrate each theme, with the participant identification number, age range and SDS score also provided for context.

**Table 1:** Participant characteristics

| Demographic | Number (%) |
| --- | --- |
| Total sample | 11 |
| Gender (identified as woman) | 11 (100) |
| Age range in years |  |
| 25-35 | 2 (9) |
| 36-40 | 2 (18) |
| 41-45 | 4 (36) |
| 46+ | 3 (27) |
| Aboriginal/Torres Strait Islander | 3 (27) |
| Sexual Orientation |  |
| Heterosexual | 7 (63) |
| Bisexual/Queer | 4 (37) |
| Relationship Status |  |
| In a relationship | 6 (55) |
| Experiencing Homelessness | 5 (45) |
| Mode of MA use |  |
| IV | 11 (100) |
| Inhalation | 2 (also smoked) |
| Frequency of use (last 30 days) |  |
| >2 times per week | 11 (100) |
| Daily | 7 (63) |
| Duration of frequent use |  |
| <5 years | 5 (45) |
| 5-10 years | 4 (37) |
| 20 years + | 2 (18) |
| Dependence on MA |  |
| SDS >= 4 | 8 (72) |
| Self-described dependence | 7 (63) |
| Reported Use/Dependence on Other Drugs |  |
| Opiates | 10 (90) / 7 (63) |
| Alcohol | 11 (100) / 1 (1) |
| Cigarettes | 10 (90) / 7 (63) |
| GHB/GBL | 8 (72) / 0 (0) |
| Cannabis | 11 (100) / 3 (27) |
| BZD | 10 (90) / 2 (18) |
| Self-reported severe harms related to MA |  |
| Physical* | 10 (90) |
| Mental** | 6 (55) |
| Social*** | 11 (100) |

\*Heart attack, cardiac stents, chest pain, bacterial endocarditis, oral decay, abscesses, staphylococcus skin infections, hepatitis, persistent cough
\*\*Psychosis, severe mood swings, depression, anxiety, paranoia, hallucinations
\*\*\*Intimate partner violence, domestic violence, violence, imprisonment, loss of custody of children, debt, gambling, poverty, homelessness

### Identity, stigma and resistance

Several participants emphasised the positive aspects of their MA use, such as how it enabled productivity, or as a form of recreation.

> *“My first experience … I found that it elevated my levels of perception and my levels of superwoman likeness … in a way that I could achieve many things in a short period of time*.*”* (Participant 3, age 31-35, SDS 1)

Nonetheless, stigmatising language was used by the participants themselves in describing MA use.

> *“The methamphetamine scene - I have never seen such putrid behaviour. I’ve never seen such vile creatures or predators or soulless people in my life and it has been truly a very disheartening experience*.*”* (Participant 3, age 31-35, SDS 1)

They also described the stigma they felt in being identifiable as someone who used drugs.

> *“I sort of feel like there’s a big tag on my head that says I’m a drug user (laughs). I don’t know but I just stand out*.*”* (Participant 1, age >50, SDS 5)

This affected their sense of belonging and identity within their families.

> *“My family judged me. They judged my loss of weight, they judged my appearance, they judged the fact that I had lost everything in my life”* (Participant 3, age 31-35, SDS 1)

The loss of custody of children in relation to their MA use and implications of this on their sense of self, was also discussed by participants.

> *“It’s not pleasant. I really need to go back to family. There are things that the ice has stopped me from doing like things that I should be doing*.*”* (Participant 6, age 41-45, SDS 9)
>
> *“No one ever gave me a pat on the back to say, you know “ Good, we recognise that you’ve made major changes because of your children” …you know I don’t want to be labelled like a bad parent*.*”* (Participant 1, age >50, SDS 5)

Participants characterised their own use as being different to problematic or dependent use.

> *“I certainly wouldn’t say that I am addicted to it…It takes a very strong personality to take a drug for three days and then stop because this drug is by far the most addictive drug out of all of them*.*”* (Participant 3, age 31-35, SDS 1)

Participants also highlighted the difference between their use with the use of others, both in terms of “controlled use” and better management of the consequences of use.

> *“I feel sorry for her…she doesn’t have that self-control that’s just the mind over matter thing*.*”* (Participant 3, age 31-35, SDS 1)
>
> *“Ah it’s just disgusting…she must probably shoot up her neck, so wherever the shooting site is…If I even see a couple of spots on me, I’m straight to the doctor. I don’t wanna turn out like that*.” (Participant 1, age >50, SDS 5)

### Interpersonal violence

Several participants spoke of their intimate partner relationships as being innately linked with their use of MA. These relationships were also associated with the experience of violence.

> *“It was in a domestic violent relationship and the deal with him was if I were to use it, I had to inject it. So I started injecting it. Before that I was smoking it. It was a control thing for him, being the control freak*.*”* (Participant 4, age 36-40, SDS 10)
>
> *“If I ever get in a co-dependant relationship or a violent relationship it was usually based around using of ice. If they went to jail I stopped using”* (Participant 6, age 41-45, SDS 9)

Partners were also described as key to the participant’s initial use of MA.

> *“I never used to like the ice…it was only through a boyfriend I was with, but before that the thought of it - I hated it. It was first it was around having sex, get on the high before having sex or while having sex. I did it at the beginning to please my partner – I never really did it for me*.*”* (Participant 4, age 36-40, SDS 10)

### Institutionalised stigma

Participants identified situations in which they faced institutional prejudice and stigma, particularly within the healthcare and criminal justice systems.

> *“With police you know, I always have issues with them you know because of my drug history. Once they know you’re a drug user they treat you different…To them I think you know you’re a piece of shit…so they talk down to you…Even in domestic violence situations where I’ve been badly bashed or whatever they still treat you the same*.*”* (Participant 4, age 36-40, SDS 10)

This extended into healthcare interactions, and participants reported that this impacted on their willingness to ask or receive help.

> *“I don’t know if I was paranoid but yeah with some doctors I felt once they realised you’re a sex worker or a drug user their whole persona changes*.*”* (Participant 4, age 36-40, SDS 10)
>
> *“I had to wait two hours… and it wasn’t until I started crying and going just because you couldn’t… you know no one can see my symptoms*.*”* (Participant 8, age 41-45, SDS 3)

Participants identified how feelings of shame affected their capacity to seek help.

> *“I feel a bit shame about what I’ve been through and I didn’t wanna talk about it cos I didn’t know how to express it. Now that I’m over it and I’ve realised what I’ve been through I can talk and express it*.” (Participant 2, age 25-30, SDS 11)

Even when seeking help, participants emphasised the challenge in communicating their distress to healthcare workers.

> *“No one knew what I was going through because they hadn’t been through it, or they never knew anyone in that situation before…I wasn’t speaking the right words*.*”* (Participant 2, age 25-30, SDS 11)

### Health service delivery preferences

We identified three key themes around participant’s preferences for health service delivery: trusted continuous care; care that’s integrated with other services; and is free of judgment.

#### Trusted continuous care

Participants valued continuity of care from healthcare professionals that knew their individual history and they had built a therapeutic relationship with over an extended period of time. It gave participants a sense of trust and belonging.

> *“Sometimes I guess I feel shy around people I don’t know. The last couple of times I’ve been shifted around to a different person and I’m not gonna start telling them my dark secrets*.*”* (Participant 7, age 41-45, SDS 11)
>
> *“I have…a problem with feeling comfortable with certain people and trust issues. If I don’t feel comfortable or if I don’t like you I won’t be sitting there. It’s taken weeks, months if not years before I even sit down and have a conversation with you*.*”* (Participant 4, age 36-40, SDS 10)

#### Integrated with other services

Participants valued services who had an integrated approach to medical care, psychological and social service support.

> *“It’s the place that I’ll go to for everything…they’re the best people…I see my psychiatrist there, my doctors there, and my counsellor…my folder’s about that thick [laughs]*.*”* (Participant 1, age >50, SDS 5)
>
> Culturally appropriate services were also valued.
>
> *“[Aboriginal Health Worker] gets what I like, what I don’t like. She even knows the clothes I like to wear…Whenever I get any of the ladies that are downstairs, that are non-Indigenous, they always come out with old people clothes*.*”* (Participant 11, age 46-50, SDS 9).

#### Non-judgmental

Participants feared experiencing discrimination due to their drug use when seeking help from health professionals, and described how non-judgemental care improved their interactions with health services.

> *“I like it because they don’t discriminate … that they really make me feel I’m not different. I can just talk to them just to let it out and I never snap. [At other places] it was like guilty I had to prove myself innocent*.*”* (Participant 1, age >50, SDS 5)

## Discussion

In this study, we explored the perspectives of a sample of 11 diverse women who regularly use MA and their healthcare service delivery preferences. Findings point toward several areas which might be addressed to promote earlier specialist treatment-seeking by women who use MA.

### Working with stigma

Effective assessment of substance use issues is key to facilitating earlier recognition of substance use disorders, and engagement with specialist treatment. The self-identification of problematic use was complicated for participants in the study by feelings of shame and the fear of experiencing discrimination. Women who use drugs may be more susceptible to feeling stigmatised than men (Meyers et al., 2021), and are reported to be more likely to experience stigma as a barrier to treatment (Green, 2006). Countering stigma is a key competency for professionals working with this group (Bielenberg, Swisher, Lembke, & Haug, 2021). Green (2006) found that women were most likely to seek help for substance use in primary healthcare and mental healthcare, highlighting the need for effective engagement in for both AOD and non-AOD services. It also suggests a role for specialist MaUD treatment programs to work with and upskill healthcare professionals in non-specialist AOD and other healthcare settings

It is also notable that participants readily identified benefits to their use of MA, such as increased productivity and sociability. People may not readily identify their ambivalence around reducing or ceasing MA use in healthcare settings, given the stigma of drug-related pleasure (Pienaar & Dilkes-Frayne, 2017). Healthcare professionals should recognise there are different stages in readiness to addressing drug use, and have a number of strategies to engage accordingly. Motivational interviewing, for instance, takes account of readiness to change, and can be effective after just one session when delivered by healthcare professionals in non-AOD settings (VanBuskirk & Wetherell, 2014).

### Working with social networks

A prominent theme among study participants was the interaction between social relationships, their experience of MA use, and treatment seeking. Intimate relationships were implicated in onset and continuation of MA use for some, and relationships involving trauma and violence were common. Women with substance use disorders have been reported as having higher rates of trauma histories than men (Simpson et al., 2016), with up to 80% of women who seek treatment for substance use disorder estimated to have lifetime histories of sexual and/or physical trauma (López-Castro, Hu, Papini, Ruglass, & Hien, 2015). This underscores the need for trauma and violence-informed care in all settings (Wathen, MacGregor, & Beyrem, 2021), with health professionals trained in the empathetic eliciting and recognition of intimate partner violence, robust systems of referral, and an understanding of the impact of trauma on health and help-seeking.

A number of the participants had experienced engagement with child protection services, consistent with substance use being recognised as a predictor of children being taken into care at birth (Wall-Wieler, Roos, Brownell, Nickel, & Chateau, 2018). An Australian study of a sample of MA smokers found that in addition to experiences of poverty and homelessness, accessing treatment was also associated with an increased likelihood of child-removal for women (Ward et al., 2021). The risk of child removal in addition to stigmatisation and high rates of disadvantage and trauma for this population (O’Connor et al., 2020; O’Connor, Harris, Hamilton, Fisher, & Sachmann, 2021) underline the need for careful assessment and adequate support for this population when engaging with services.

Importantly, relationships were regarded as resource and a driver for treatment seeking and utilisation by participants. This mirrors the growing recognition of the role of significant others (Ariss & Fairbairn, 2020) and social networks as a form of recovery capital (Best, Vanderplasschen, & Nisic, 2020; Collinson & Hall, 2021), and the inclusion of family and friends in treatment planning presents an opportunity to enhance outcomes in MaUD treatment. Further research is required to build the evidence base for family and social network inclusive practices in MaUD treatment.

### Countering power imbalances

Participants reported that experiences of stigma in interactions with healthcare and other institutional settings were common. This led to caution around disclosing their MA use in case it led to discriminatory attitudes or behaviours towards them when they were especially vulnerable, and underpins the value participants held for trusted relationships built over time with individual professionals in those institutions. Hospitals and other healthcare settings can be experienced as unsafe spaces by people who use drugs (McNeil, Small, Wood, & Kerr, 2014; Pauly, McCall, Browne, Parker, & Mollison, 2015). The inclusion of workers with lived experience in healthcare teams can be an effective strategy in reducing fear of discrimination and empowering clients (Olding, Cook, Austin, & Boyd, 2022). People may feel also more comfortable receiving healthcare outside of institutional settings, and so the delivery of specialist MaUD treatment through outreach and community partnerships, and with technological innovations such as telehealth, may also be of value in addressing these provider-client power imbalances.

### Integrated and structurally competent service delivery

Participants in the study had needs across multiple domains, encompassing physical and mental health needs, income support, social support and justice involvement. This need for integrated care to support the whole of a person’s health needs through effective working across healthcare and other services has also been highlighted by other studies with people with substance use disorders (Petzold et al., 2021; Savic, Best, Manning, & Lubman, 2017).

The participants’ accounts of disadvantage are also better understood as resilience in the face of structural vulnerability (Bourgois, Holmes, Sue, & Quesada, 2017), rather than behaviours or social risk factors. Such an understanding focuses on the ways in which social structures and power relationships make specific groups more or less likely to develop a substance use disorddr, which is then compounded by inequities in access to healthcare treatment. As McKenna notes, poor and minority women who use drugs exist in risk environments characterised by multiple levels of structural, physical and symbolic violence (McKenna, 2014). This vulnerability is reinforced by stigmatizing drug policies, laws and media portrayals (McKenna, 2014). The multiple axes of structural vulnerability recounted by the participants in this study, not just gender, but also race, and poverty, underscore the need an intersectional approach to planning MaUD treatment services. Structurally competent healthcare (Metzl & Hansen, 2014) seeks to produce workers and systems who recognise the effects of structural disadvantages on health and develop responses that prevent further harms. It is especially relevant in addressing the challenges to health equity that arise from systemic discrimination for people who use drugs (McNeil, Kerr, Pauly, Wood, & Small, 2016; Treloar et al., 2021), as well as inequities arising due to gender (Mackenzie, Gannon, Stanley, Cosgrove, & Feder, 2019; Preis, Garry, Herrera, Garretto, & Lobel, 2020).

### Limitations

Our study is subject to a number of limitations. Firstly, the size and source of our sample means that the study should not be taken as a representative sample of women who use MA. For instance, the sample was drawn from those already accessing public health services, so may have missed more marginalised women. Conversely, those accessing privately funded services may also have been missed. We also did not explicitly seek individuals who identified as transgender or gender diverse. This necessarily limits the generalisability of this study.

## Conclusion

This study explored the perspectives of a diverse group of women on their MA use and preferences for healthcare. Findings show a complex interaction between the management of identity, self-stigma and the fear of discrimination and showed personal relationships as being both a challenge and a resource. Healthcare institutions were not seen as safe spaces, with integration and continuity of care delivered by non-judgemental staff valued by the participants, and the need for trauma and violence informed care in all settings was clear. Findings may also have relevance for healthcare interventions for a range of other substance use disorders. Future research on effective anti-stigma interventions for healthcare professionals, family inclusive practice, and structurally competent care will be of value in providing better access to MaUD treatment services for women.

## Data Availability

Ethics approval does not permit sharing of the data.

## Acknowledgements

The authors would like to acknowledge and pay their respects to the custodians of the land where the project was undertaken, the Gadigal people of the Eora Nation. They also offer their sincere gratitude to the women who participated in the study and shared their stories. Thanks to Leanne McQuiston for her support with transcription.

